# RANDOM NUMBER GENERATION: A ANALYSIS OF BENFORD`S LAW IN UNDERGRADS STUDENTS AND GERIATRICS PATIENTS

**DOI:** 10.1101/2024.08.17.24312153

**Authors:** Moacir Fernandes de Godoy, Pedro Henrique Alves de Freitas Martins

## Abstract

**Objectives:** Study the correlation between RNG by human cognition and Benford’s Law in undergraduate students at a Medical School in the countryside of the state of São Paulo and geriatric clinic patients.

**Design:** The study collected data from the students and patients from the Robust Elderly Clinic of a tertiary Hospital between August 2022 and July 2023. Data collection involved a questionnaire on age, gender, education, ethnicity, occupation, and a table with 5 rows and 10 columns for the insertion of 50 numbers, chosen by the participant.

**Results:** A total of 263 forms were collected. The average age was 27.10 (IQR- 3) years, with 66.5% being female. Frequencies of the first significant digit were: 25.59% for 1; 15.35% for 2; 10.98% for 3; 8.65% for 4; 9.67% for 5; 7.02% for 6; 8.22% for 7; 6.81% for 8, and 7.70% for 9. Applying the Chi-Square test, no statistically significant difference was found (critical χ ^2^ 15.507; obtained χ ^2^ 5.36). Applying Pearson’s Coefficient, the value of r was 0.98. Using the Euclidean distance, the P-value was 0.9284.

**Conclusion:** A high correlation between RNG by the human mind, in students and Robust Elderly patients, and Benford’s Law was detected.

## Introduction

### Random number generation

The process of Random number generation by the human mind has been a site for investigation for quite some time. According to Pernaud N^1^, It was observed that even as it can create true Random sequences, the human mind tends to generate pseudorandom numbers circles that based on the repetition of some patterns and suppression of others create cognitive fingerprints in the Generation, as seen by Schulz et all^2^. In other words, human cognition does not always produce true Random phenomena.

Random number generation happens based on the relation of the left dorsolateral prefrontal cortex region and the superior temporal cortex, the last being associated with sequential counting, there being a negative modulation between the two cortexes, in which the first inhibits the second. Jahanshahi et all^3^ found this through the reduction of the blood flow in the counting region as the dorsolateral region is activated.

Working memory, as described by Baddeley in 1974, emerges as a model to explain the cognitive capacity to retain multifaceted information—visual, spatial, and temporal—and perform mental operations on them within a short period. Working memory should be regarded as a brain network not localized in a single brain region, but rather as a property that arises from the functional relationship between the prefrontal cortex (PFC) and the rest of the brain, as stated by D’Esposito M. Additionally, Persaud N demonstrates how changes in working memory, self-control, and problem-solving are reflected in RNG tests.

Therefore, it is observed that variance in the human executive function: such as work memory, self-control, reasoning, and problem-solving, are reflected in the cognitive capability of creating true randomness, according to Pernaud N^1^. Based on this premise, the Random number generation process has been used to investigate diseases such as Parkinson’s, as shown by Law et all^5^. At the same time, another phenomenon has intrigued mathematicians Around the world, Benford’s Law.

### Benford’s Law

In 1881, Simon Newcomb saw that in a logarithm book that came into his possession, there were distinct signs of greater usage of the initial pages, with the higher-numbered pages experiencing less utilization.

This same observation was also made by Frank Benford, an American physicist who, after analyzing over 20,000 samples from various types, such as river lengths, addresses, sports statistics, population sizes, and physical constants, found that instead of a uniform distribution of initial significant digits (i.e., the leftmost digits of values excluding zero), there was a distinctive prevalence following a logarithmic scale showed Benford F^6^.

The analyses conducted by Newcomb and Benford led to the conclusion that the initial significant digits of certain numerical series did not occur uniformly but rather exhibited a non- uniform distribution among the numerals 1 through 9.

Benford’s Law has been utilized for various purposes, including fraud detection in academic articles, according to Horton J et al^8^, public procurement audits, according to da Hora Sampaio A et al^9^, and international trade, as described by Cerioli A et all^10^. This detection is based on the fact that artificially fabricated data by individuals tends to deviate from the law, as shown by Gauvrit NG et al^11^

It is important to note, as pointed out by Cymrot R et al^12^, however, that for the analysis to be feasible, the data must exhibit similar phenomena, featuring independent data points within relatively large samples from unmanipulated sources. In other words, data with upper and lower bounds, such as average salaries or population heights, do not adhere to Benford’s Law, nor are numbers subject to active human manipulation. Natural numbers, on the other hand, such as mortality rates and birth rates, follow the distribution as shown by Benford F and Newcomb S.^6,7^

### Benford in healthcare

Within the realm of healthcare, Benford’s Law holds the potential to serve as an inexpensive and easily reproducible screening tool. However, the application of the Law in this field is not yet widely explored.

There is a dearth of studies evaluating whether human-generated data aligns with this distribution. Hsu E.H and Kubovy DA^13,14^, in their initial studies addressing this possibility, required participants to write small series of 4-6 digits that were supposed to be original, all yielding negative results in terms of Law conformity, with at most a hint of the influencers’ impact on the participants.

Nevertheless, this consensus of non-conformity shifted in the early part of the last decade, as Diekmann A^15^ utilizing regression patterns and multiple-digit repetition pointed towards a Benfordian distribution. Alongside this, Burns^16^ created the concept that the key question is not the exact adherence to Benford’s Law in numbers generated by individuals, but rather the extent to which a Benfordian bias, a distortion of the spontaneous numerical pattern generated by the human mind that approximates Benford’s Law, would be significant and prevalent in spontaneous numerical Generation. It is important to address that, the Law has never been used as a screening heath test before.

Therefore, this study aims to explore the correlation between spontaneous numerical generation by human cognition and Benford’s Law among undergraduate students, as well as among Robust Elderly patients at a geriatric outpatient clinic, and to comprehend the potential presence of a Benfordian bias within this population.

## Methods

### Sample characterization

Upon approval from the appropriate Ethics Committee, this quantitative observational analytical cross-sectional prevalence study was conducted with a convenience sample. Data collection took place among students in the 1st to 4th year of the Medicine program, 1st and 2nd year of the Nursing program, and 1st to 3rd year of the Psychology program, as well as with patients from the Robust Elderly Outpatient Clinic within the Geriatrics department at the Hospital. The data collection period extended from August 2022 to July 2023.

Participants were divided into two major groups: university students and patients from the Geriatrics department. This division aimed to form the largest and most diverse group of individuals for data collection. Exclusion criteria encompassed failure to provide informed consent, age below 18 years, mild or moderate cognitive impairment reported by the primary caregiver, and incomplete completion of the data collection materials. Additionally, the classification of robust elderly individuals and the cognitive assessment for mild or moderate cognitive impairment were carried out by the geriatrics department before the study, considering factors such as sarcopenia, grip strength, gait speed, ability to sit and rise without assistance, weight loss, and feelings of exhaustion. Those classified as frail or pre-frail were not included in the study.

Regarding sample size, as indicated in the literature, conformity with Benford’s Law requires a minimum sample size of 50-100 observations, as shown by Kenny DA^15^, with no maximum limit specified. Consequently, the total student body was summed up, resulting in 500 students (320 from medicine, 60 from psychology, and 120 from nursing). An estimate provided by the Geriatrics department at the school Hospital projected a frequency of 40 patients attending the outpatient clinic. A sample size of 50% of the observed population was determined, yielding approximately 270 participants.

### Instruments and data collection

For data collection, a two-part form was employed. The first part consisted of a questionnaire, encompassing inquiries regarding age, gender, educational level, ethnicity, and occupation. The second part comprised a table with 5 rows and 10 columns, providing spaces for the insertion of 50 spontaneously generated numbers by each individual.

During data collection, participants were informed about the research purpose, risks, contributions, and the option to withdraw at any point. They were then provided with the questionnaire. Following this, before the application of the questionnaire, the Informed Consent Form was presented. If, after reading and explanation, participants agreed to continue the research, the process was initiated.

If the patient was illiterate, the study and informed consent forms were read and presented to their primary caregiver, after which participation in the study was discussed between them. If the patient was illiterate and did not have a primary caregiver, they were disqualified from participating due to the inability to complete the informed consent form. For student participants, the form was given for self-completion. For those in the Geriatrics service, the researcher conducted the questionnaire. In the latter case, to collect the 50 digits, the researcher, at near one-second intervals, prompted participants to respond with a number of their choice. Participants were informed that the chosen numbers had no limitations and could encompass various decimal places, be below zero, include decimals, or even negative numbers. If a number below 1 were chosen the first digit that was not zero would be the one considered in the analysis. (Ex: If 0,45 is chosen, then the first digit is 4).

### Statistical analysis

Upon data collection, the data was tabulated in a Microsoft Excel spreadsheet to create a database containing variables such as gender, age, ethnicity, educational level, occupation, and the first significant digit (1-9) of the 50 spontaneously generated numbers by each participant. These digits were then compared with the expected values according to Benford’s Law.

Descriptive statistical analysis of qualitative data was carried out by calculating the frequency for age, gender, ethnicity, and occupation, as well as frequency counts for the significant digits. Formulas in the spreadsheet initially calculated the sum of the first digits for each individual. Subsequently, percentages of each digit were calculated in relation to the total count for each participant.

For inferential statistical analysis of qualitative variables, two different statistical tests were employed. Firstly, the most commonly used test within the Benford literature, is the Chi-Square (χ^2^) test, a hypothesis test that compares proportions, examining discrepancies between observed and expected frequencies for a given event. Additionally, the Pearson correlation test was used. Correlation coefficients (r) were classified as follows:

- r = 0.10 to 0.30 (weak)
- r = 0.40 to 0.60 (moderate)
- r = 0.70 to 1 (strong)

Moreover, for a quantitative analysis of the Benford variable, the test of cumulative distribution based on Euclidean distance was employed. This test, adapted by Campanelli^18^ for the Law, allows for the calculation of the p-value and the assessment of compliance with Benford’s Law.

The null hypothesis (H0) was that the observed distribution of the first significant digit would be the same as the expected quantity (logarithmic proportion), while the alternative hypothesis (H1) was that the observed distribution would differ from the logarithmic proportion. Furthermore, a significance level of P ≤ 0.05 was considered statistically significant.

## Results

In 9 months of data collection, a total of 263 forms were collected. For analyzing age, the participants were divided into two subgroups, the Elderly one, with an average age of 78,11 years, an interquartile range (IQR) of 11 (Q1 of 72 and Q3 of 83), and a standard deviation of 6,68. The Not Elderly, with an average age of 21,7 years and 236 participants, an interquartile range (IQR) of 3 (Q1 of 20 and Q3 of 23), notice that the data did not follow a normal distribution, with 66.5% (175 participants) identifying as female. Regarding ethnicity, 86.3% of the participants self-identified as white; 4.6% as Asian; and 9,1% as black and Brown. In terms of educational attainment, all the members of the Not Elderly subgroup had attained a higher education. On the contrary, in the Elderly group, 8% had completed a higher education; 12% had completed high school; 62% had only completed elementary school; and 19% were illiterate.

Concerning the first significant digits, following data tabulation, the following results were obtained:

For the analysis of conformity, we compared the above numbers with the expected proportions of each of the nine responses (1-9), according to Benford’s Law, applying Chi- Square tests and Pearson’s Linear Correlation. The results of these analyses are presented below:

Applying the Chi-Square (χ^2^) Test yielded a result of 5.36, indicating the absence of statistically significant differences between the data obtained from the overall participant count and Benford’s Law, considering a significance level of 0.05 and a Critical χ^2^ of 15.507.

Continuing the analysis, applying Pearson’s Linear Correlation Coefficient to the data obtained in Table 1, a correlation of 0.98 was observed, signifying a positive and strong correlation.

**Table 1.** Benford Comparison and 1st significant digit general Count.

| 1st Significant Digit | General Count | % Total | % Benford |
| --- | --- | --- | --- |
| 1 | 3365 | 25.59 | 30.10 |
| 2 | 2018 | 15.35 | 17.61 |
| 3 | 1444 | 10.98 | 12.49 |
| 4 | 1138 | 8.65 | 9.69 |
| 5 | 1272 | 9.67 | 7.92 |
| 6 | 923 | 7.02 | 6.69 |
| 7 | 1081 | 8.22 | 5.80 |
| 8 | 896 | 6.81 | 5.12 |
| 9 | 1013 | 7.70 | 4.58 |

Utilizing the adapted Euclidean distance test, which allows for the calculation of the confidence value (p) for the sample in question, a calculated (p) value of 0.93 indicated conformity between the collected data and Benford’s Law.

In line with the findings, when isolating the geriatric age group, aged 65 and older, we had 26 interviewees, with an average age of 78.12 years. Following the count, the following results were obtained:

Applying the Chi-Square Test (X2), we obtained a result of 5.47, indicating the absence of statistically significant differences between the data found in the overall participant count and Benford’s Law, considering a significance level of 0.05, with a Critical X2 of 15.507.

Continuing the analysis, when applying Pearson’s Linear Correlation Coefficient to the data obtained in Figure 2, we observed a correlation of 0.98, which is considered a strong, positive correlation.

**Figure 1.**
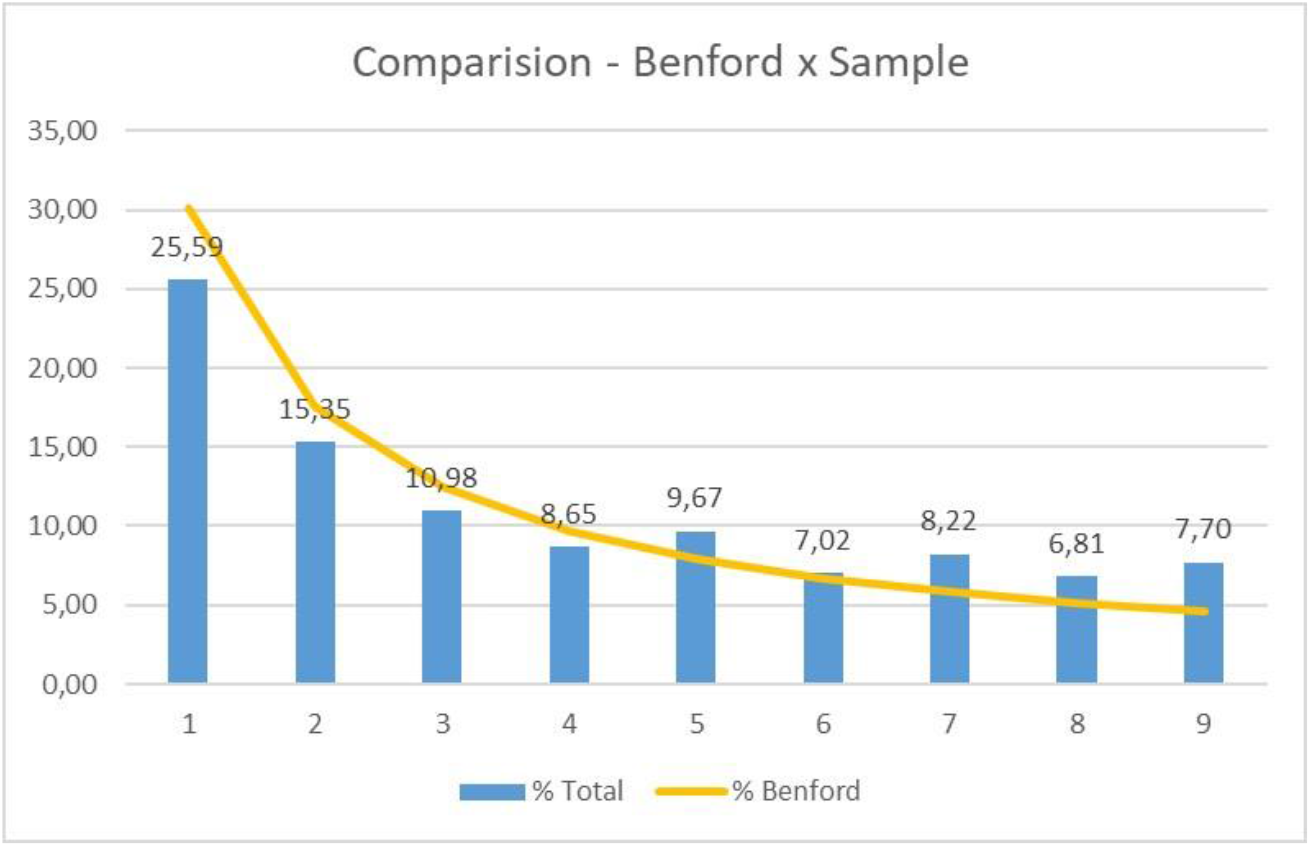
General Count

**Figure 2.**
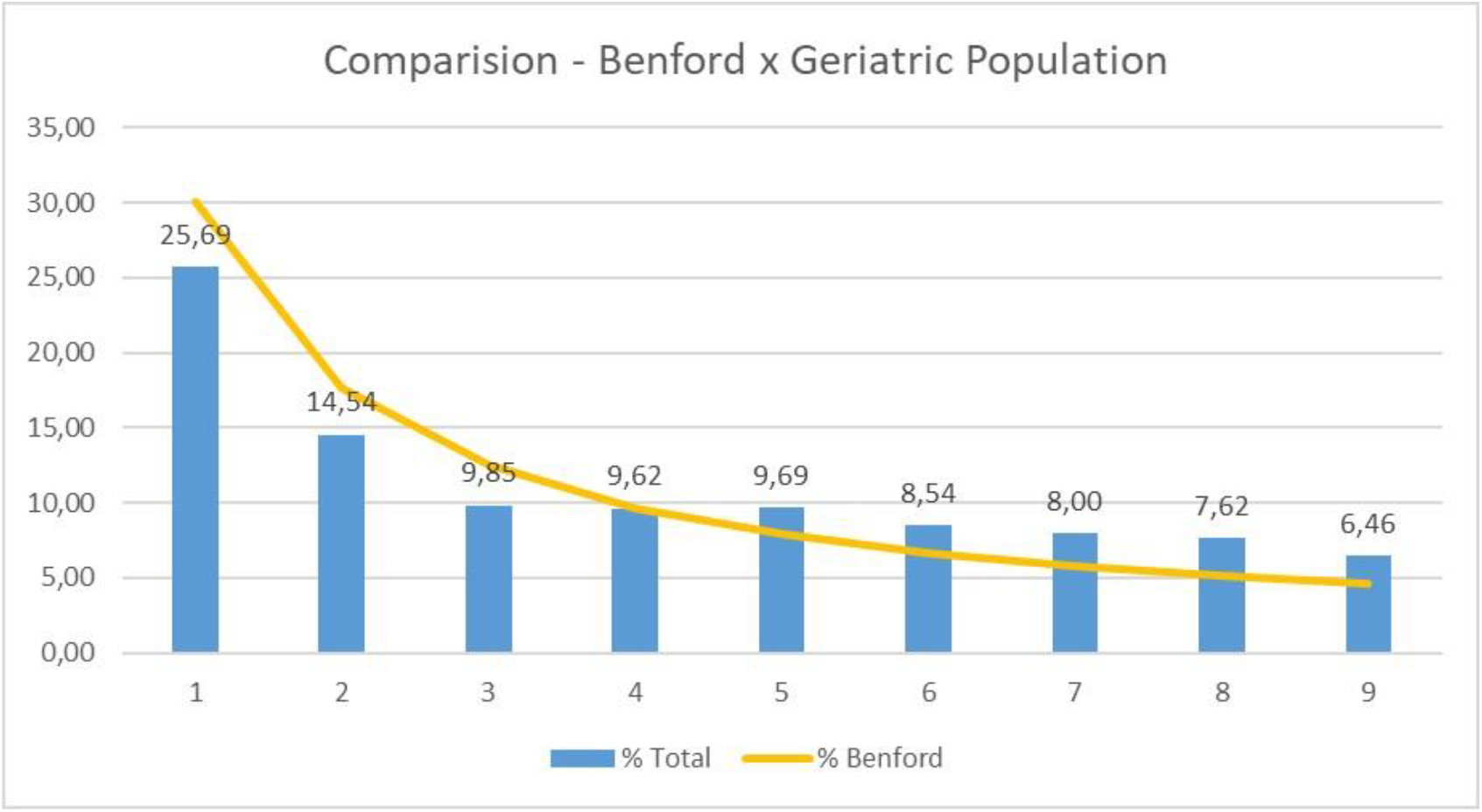
Benford X Geriatric Population

Using the adapted Euclidean Distance test, which allows the calculation of the confidence value (p) for the sample in question, we calculated a (p) of 0.92, indicating conformity between the obtained data and Benford’s Law.

Regarding the second significant digit, following tabulation, the following results were obtained:

Applying the Chi-Square (χ^2^) test to the data in Figure 3 yielded a value of 20.70, indicating statistically significant differences between the data obtained from the general count of the second significant digit and Benford’s Law, considering a significance level of 0.05 and a Critical χ^2^ of 15.507.

**Figure 3.**
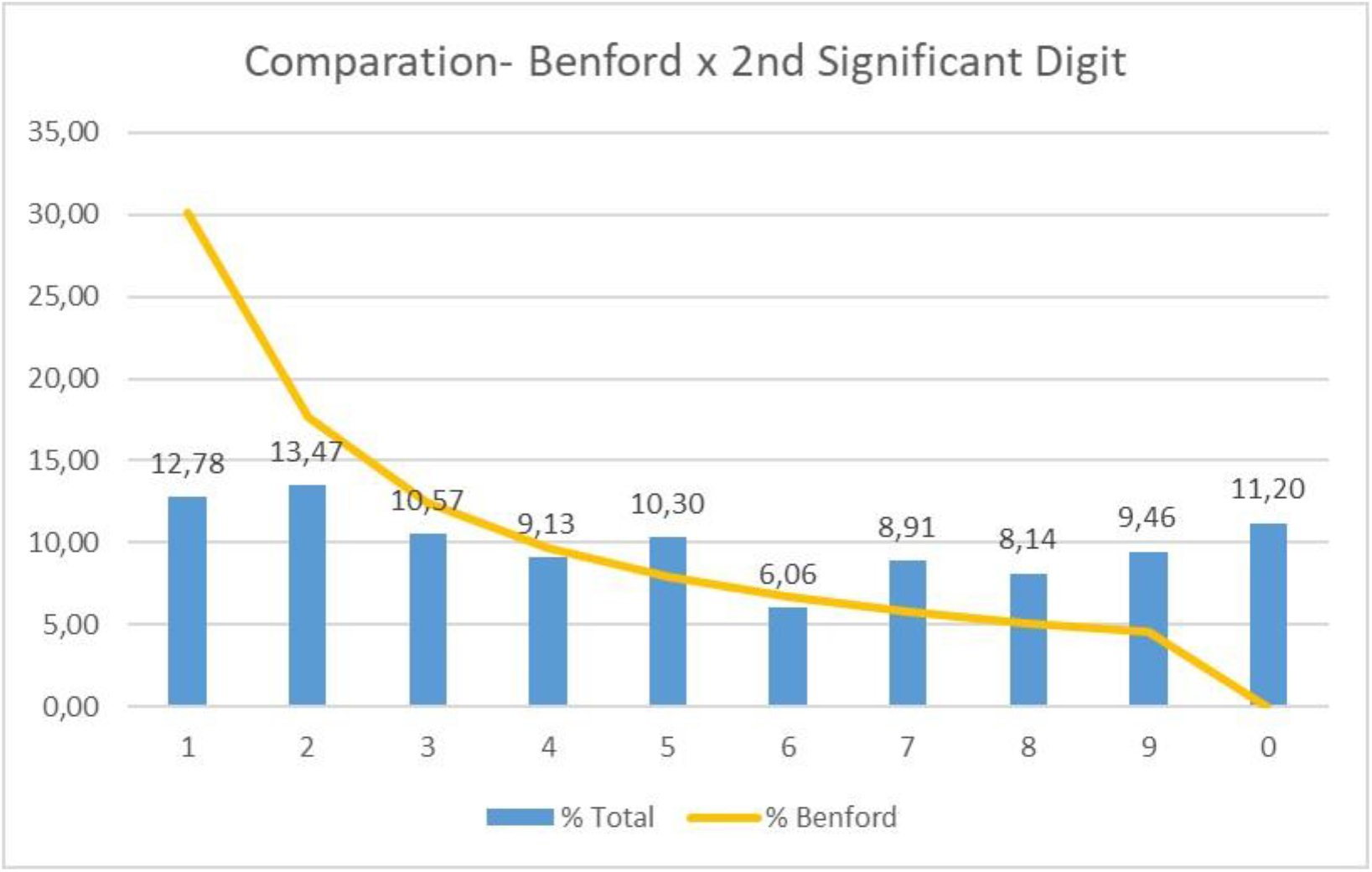
2nd Significant Digit and General Count

## Discussion

Regarding the obtained results, it was observed that voluntarily generated numbers by the participants tended to Benford’s Law in their first significant digit. This finding contradicts the initial studies that demonstrated a fit of human cognition to the Law. These early studies, developed by Hsu, Hill, and Kubovy ^12,14,19^, instructed participants to create a single number with four decimal places arbitrarily, resulting in a pattern of uniform distribution. However, this pattern has been contested in recent decades when alternative methods utilizing non-arbitrary numbers, as showed by Diekmann^14^, and a larger quantity of numbers related to real data, such as mortality rates and river sizes, according to Burns BD^20^, revealed a distortion in the direction of the Law.

Similarly, our study, by instructing participants to generate 50 numbers, aligns with recent findings, indicating that even with arbitrary numbers, a high quantity of generated numbers would exhibit the Benfordian pattern. This research follows the line explored by Burns^20^, suggesting a shift in perspective regarding the fit of human cognition to the Law. Burns proposes that seeking an exact fit, as pursued in initial studies, oversimplifies the multifaceted processes involved in spontaneous numerical generation. Instead, it’s more relevant to explore the presence of a Benfordian bias – the extent to which human cognition displays a preference for lower digits in the process of numerical generation. This perspective is also congruent with the elevation of the digit 5, which is in line with the Benford pattern, according to Scott et al and Burns & Krygier^16,21^.

In this context, our study demonstrates how such a bias can be expressed through an easy and cost-effective screening tool, with high applicability and reproducibility.

When exploring the Benfordian bias, several hypotheses arise. The Recognition Hypothesis, according to Chi D & Burns B^22^, suggests that if the world is comprised of information that adheres to Benford’s Law, individuals will become sensitive to this statistical situation. In essence, the decision-making process of these individuals would be conditioned by implicit knowledge of the Law. However, this hypothesis has been demonstrated to be untrue by Tipodi M and Chi D & Burns B^22,23^.

Consequently, the Integration Hypothesis, according to Berger A., & Hill, T^24^, suggests that the emergence of Benford’s Law is a product of the cognitive process of numerical combination during spontaneous numerical generation. In other words, Benford’s Law will be expressed through cognition as a result of the intrinsic processes of numerical combination performed by our brain when generating numbers spontaneously; in other words, it will be a reflection of a cognitive process. This brings the focus back to the process of spontaneous numerical generation, which according to Hoshi Y et all^25^ and Jahanshahi M^4^ et al, is based on the left dorsolateral prefrontal cortex (DLPFC) and the superior temporal cortex with more intense activation in the ventrolateral region of the DLPFC, according to o Agbangla et al^26^. Simultaneously, the connection between the DLPFC and the rest of the brain gives rise to working memory, as shown by D’Esposito M^4^, which serves as a model to explain the cognitive ability to retain multifaceted information and perform mental operations on it, according to Baddeley A^3^ and Baddeley A & Hitch G^,27^. Notably, spontaneous numerical generation, as demonstrated in this study to exhibit a Benfordian bias, is used as a means to evaluate executive functions and working memory, according to Persaud N.^1^.

Thus, a potential link between the emergence of this Benfordian bias and the same cognitive processes related to executive functions and working memory is observed. This raises the possibility that this bias, through spontaneous numerical generation, could act as an indirect measure of executive functions and working memory. The study’s main finding is the capacity to produce the Benfordian bias using the presented instruments.

It is important to emphasize that the presence of Benford bias in human cognition occurs when utilizing RNG techniques. Thus, other cognitive processes, such as the formation of fraudulent data, utilize different cognitive areas, as demonstrated above. Consequently, since these are different cognitive processes in different locations, Benford’s Law can concurrently be used to investigate fraudulent data formed by human cognition and potentially analyze working memory through the Benfordian tendency of cognition, as demonstrated by RNG. Therefore, the study focused on demonstrating how the present data collection instrument is capable of expressing the Benford bias under RNG conditions. Consequently, the analysis of individuals was conducted in subgroups rather than separately, due to the primary objective of the study: to establish the method in question, with the subsequent evaluation of its utilization as a cognitive screening test.

The main limitations of the study are related to the participant pool, which although diverse in age and substantial in number, is somewhat skewed towards young individuals in higher education. Additionally, despite various advances in the statistical field related to assessing the Benfordian bias, some methodological instability persists due to the inherent characteristics of Benford’s Law.

In conclusion, revisiting the study’s objective of analyzing the correlation between human cognition and the Benfordian bias in a specific, it is evident that such a correlation has been confirmed. Additionally, the feasibility of utilizing the proposed data collection method to assess the Benfordian bias in human cognition has been demonstrated, with low cost and a large potential for clinical application.

## Data Availability

All data produced in the present study are available upon reasonable request to the authors

## Acknowledgments

The authors declare the submission, evaluation, and approval of this study by the relevant ethics committee. Furthermore, they reaffirm their commitment to providing any requested data upon reasonable request. The authors declare no significant conflit of interest.

